# Smoking trajectories over the first year of the pandemic in UK middle-aged adults: evidence from the UKHLS COVID-19 study

**DOI:** 10.1101/2022.05.11.22274937

**Authors:** Thierry Gagné

**Affiliations:** Research Department of Epidemiology and Public Health, University College London, United Kingdom; International Centre for Lifecourse Studies in Society and Health, United Kingdom

## Abstract

**Background:** The COVID-19 pandemic has altered the conditions leading people to smoke. Multiple studies have examined changes in population levels of smoking at the start of the pandemic. However, conclusions remain mixed due to the high proportion of studies with poor methods and short follow-up periods.

**Methods:** This study used longitudinal data from the UKHLS COVID-19 study to derive smoking trajectories among 4,130 UK adults aged 35-64 across four time points over the first year of the pandemic (2018-19, April 2020, September 2020, and January 2021). Random-effects models were used to examine subject-specific changes across time points.

**Results:** Between the pre-pandemic estimate and January 2021, there was a significant decline in smoking from 14.8% to 13.1% (PR = 0.89, 95%CI 0.83-0.95). The number of cigarettes smoked per day among smokers increased in April 2020 (*B* = 0.5, 95%CI 0.0, 1.0) and September 2020 (*B* = 1.0, 95%CI 0.4, 1.5), but declined back to pre-pandemic levels in January 2021 (*B* = 0.3, 95%CI -0.3, 0.8). These changes did not vary by sex, ethnicity, relationship status, education, occupation, or household income.

**Conclusion:** Among UK adults aged 35-64, there has been a slight decrease in smoking which was maintained up to January 2021. Whereas there was an increase in cigarette consumption among smokers at the start of the pandemic, this was no longer observable in January 2021. The findings support the argument that the first year of the pandemic is unlikely to have had a negative effect of most middle-aged adult smokers’ trajectory.

## 1. INTRODUCTION

The COVID-19 pandemic dramatically altered the conditions leading people to smoke in the UK. While a large number of studies have been published to understand changes in population levels of smoking over the past two years, evidence is mixed on whether it has actually increased, decreased, or stayed the same over time.^1–3^ A recent meta-analysis of early changes estimated both a decrease in smoking prevalence and an increase in cigarette consumption among smokers.^2^ The pandemic influenced smoking in both negative (e.g., boredom, financial stress) and positive (e.g., fear of complications) ways.^1^ Some also argued that while intentions to quit smoking increased at the start of the pandemic, attempts have been less succesful because they were more likely to be unaided.^1^ Limiting our understanding of changes over time, a majority of studies used relatively poor designs, i.e., cross-sectional samples, retrospective measures, and non-representative sampling, to examine these changes.^2^ Finally, few studies have estimated whether changes at the start of the pandemic have been maintained after the end of summer 2020.^1^

To help us better understand more recent changes in smoking, we may leverage the UK cohort datasets that collected data from their cohort members during the pandemic. Among them, the UK Household Longitudinal Study (UKHLS) offers a key opportunity to derive smoking trajectories. To the best of our knowledge, only one study previously examined changes in smoking using the UKHLS. Compared with a pre-pandemic estimate in 2017-18, the authors found that the risk of smoking in UK adults aged 16+ had declined by 11% in April 2020, that this decline did not signficantly differed by age group, gender, ethnicity, or education, and that it may have been driven by cessation among lighter smokers. This brief report contributes to the current evidence gap by examining smoking trajectories in the longitudinal sub-sample of UKHLS participants aged 35-64 across the four time points where smoking was measured: 2018-19, April 2020, September 2020, and January 2021.

## 2. METHODS

### Data

The UKHLS is a nationally representative survey of the UK population following a household sample every year since 2009-10.^4^ At the start of the pandemic, it invited participants who responded in Waves 8 and 9 (2016-17 and 2017-18) to fill a total of eight surveys between April 2020 and March 2021 to better understand the impact of the pandemic.^5^ Smoking behaviour was measured three times in April 2020 (C1), September 2020 (C5), and January 2021 (C7).

This study focuses on the subsample of 4,310 participants: 1) aged 35-64, 2) with a full interview in Wave 10 (2018-19) as the pre-pandemic baseline, and 3) who responded in all COVID-19 survey waves up to January 2021. Whereas the COVID-19 substudy invited all UKHLS participants aged 16+, I focused on those aged 35-64 because non-response bias appeared to be much stronger in those aged 16-34 and 65+ when comparing smoking prevalence before the pandemic with estimates from other sources (see Supplementary Table 1).

Ethical approval was not necessary for the data analysis of this dataset.

### Measures

Smoking status (Yes/No) was measured asking “Do you smoke cigarettes?”. Cigarettes smoked per day among smokers was measured asking “Approximately how many cigarettes a day do you usually smoke, including those you roll yourself? If it varies, tell us the daily average over the last week. If it is less than 1 per day, please just enter 0.” To remove outliers, the number of cigarettes was recoded to a maximum of 40 (<1% of observations). Missingness across waves varied from 0.0% to 1.6% for smoking status and from 0.3% to 2.1% for cigarettes smoked per day.

The following variables in Wave 10 (2018-19) were considered to explore differences in changes over time: 1) sex (male / female), 2) living with a partner (yes / no), 3) having a White UK ethnicity (yes / no), 4) having a university degree (yes / no), 5) their current occupation level based on the 3-category National Statistics Socioeconomic Classification (professional or managerial / intermediate / routine or manual / not currently employed), and 6) their net household monthly income (quintile-based).

### Analysis

I first present the distribution of smoking prevalence and the mean number of cigarettes smoked per day among smokers across time points (Table 1). I then test population-average estimates (i.e., pooled Poisson and linear models with clustered standard errors) and subject-specific estimates (i.e., random-intercept Poisson and linear models) to assess changes over time in: 1) smoking prevalence among the full sample of observations (Table 2), and 2) cigarettes smoked per day among the subsample of observations that smoked (Table 3).^6^ Finally, I tested interactions and their statistical significance to see if changes varied across groups. Results are fully detailed in Supplementary Tables 2-5. All estimates were produced using the UKHLS longitudinal weight, primary sampling unit, and strata variables in Stata 17.

**TABLE 1.** Sample characteristics, UK adults ages 35-64. UKHLS COVID CW7 longitudinal sample (2018-21).

|  | **Wave 10**<br>**2018-19** | **Wave C1**<br>**Apr 2020** | **Wave C5**<br>**Sep 2020** | **Wave C7**<br>**Jan 2021** |
| --- | --- | --- | --- | --- |
|  | **%** | **%** | **%** | **%** |
| **Prevalence (%)** |  |  |  |  |
| Overall | 14.8 | 13.8 | 14.0 | 13.1 |
| Males | 15.5 | 15.1 | 15.6 | 14.2 |
| Females | 14.1 | 12.7 | 12.5 | 12.0 |
| **Cigarettes per day among smokers (mean)** |  |  |  |  |
| Overall | 14.5 | 15.7 | 15.2 | 15.0 |
| Males | 15.4 | 16.3 | 16.4 | 16.3 |
| Females | 13.7 | 15.0 | 13.9 | 13.7 |

Sample is those aged 35-64 with a full interview in the UKHLS main wave 10 (2018-19) and a valid longitudinal weight value in the COVID-19 survey wave 7 (January 2021). Estimates are weighted using the longitudinal C7 weight.

**TABLE 2.** Testing changes in smoking prevalence over time, UK adults ages 35-64. UKHLS COVID CW7 longitudinal sample (2018-21).

|  | PR | 95%CI | *p* |
| --- | --- | --- | --- |
|  | **Population-average**<br>**(Pooled with clustered SEs)** |  |  |
| **Time** |  |  |  |
| (ref. 2018-19) | --- | --- | --- |
| CW1 – April 2020 | 0.94 | 0.87-1.01 | .081 |
| CW5 – Sept 2020 | 0.95 | 0.88-1.02 | .158 |
| CW7 – Jan 2021 | **0.89** | **0.80-0.99** | **.027** |
|  | **Subject-specific**<br>**(Random-intercept modelling)** |  |  |
| **Time** |  |  |  |
| (ref. 2018-19) | --- | --- | --- |
| CW1 – April 2020 | **0.93** | **0.87-0.99** | **.026** |
| CW5 – Sept 2020 | **0.92** | **0.86-0.98** | **.017** |
| CW7 – Jan 2021 | **0.89** | **0.83-0.95** | **.001** |

*N* observations = 17,120. Sample is those aged 35-64 with a full interview in the UKHLS main wave 10 (2018-19) and with a valid longitudinal weight value in the COVID-19 survey wave 7 (January 2021). Estimates are weighted.

**TABLE 3.** Testing changes in cigarettes per day among smokers over time, UK adults ages 35-64. UKHLS COVID CW7 longitudinal sample (2018-21).

|  | **B** | **95%CI** | ***p*** |
| --- | --- | --- | --- |
|  | **Population-average**<br>**(Pooled with clustered SEs)** |  |  |
| **Time** |  |  |  |
| (ref. 2018-19) | --- | --- | --- |
| CW1 – April 2020 | 1.16 | 0.04, 2.27 | .043 |
| CW5 – Sept 2020 | 0.74 | -0.46, 1.94 | .225 |
| CW7 – Jan 2021 | 0.50 | -0.52, 1.51 | .338 |
|  | **Subject-specific**<br>**(Random-intercept modelling)** |  |  |
| **Time** |  |  |  |
| (ref. 2018-19) | --- | --- | --- |
| CW1 – April 2020 | **0.51** | **0.03, 1.00** | **.038** |
| CW5 – Sept 2020 | **0.98** | **0.42, 1.54** | **.001** |
| CW7 – Jan 2021 | 0.28 | -0.27, 0.83 | .316 |

*N* observations = 1,352. Sample is those aged 35-64 with a full interview in the UKHLS main wave 10 (2018-19) and with a valid longitudinal weight value in the COVID-19 survey wave 7 (January 2021). Estimates are weighted.

## 3. RESULTS

Smoking prevalence declined from 14.8% in 2018-19 to 13.8% in April 2020, 14.0% in September 2020, and 13.1% in January 2021. In the subject-specific model, this resulted in a 11% lower risk of smoking (95%CI 0.83-0.95) in January 2021 compared with the pre-pandemic estimate.

Transitions between smoking statuses suggest that most of the changes occurred by April 2020. For transitions among smokers: 1) 16.1% of smokers in 2018-19 were non-smokers in April 2020; 2) 7.4% of smokers in April 2020 were non-smokers in September 2020, and 3) 11.1% of smokers in September 2020 were non-smokers in January 2021. For transitions among non-smokers: 1) 1.7% of non-smokers in 2018-19 were smokers in April 2020; 2) 1.3% of non-smokers in April 2020 were smokers in September 2020, and 3) 0.8% of non-smokers in September 2020 were smokers in January 2021.

The mean number of cigarettes smoked per day among smokers varied from 14.5 in 2018-19, 15.7 in April 2020, 15.2 in September 2020, and 15.0 in January 2021. In the subject-specific model, this resulted in a 0.51 increase in April 2020 (95% 0.03, 1.00), a 0.98 increase in September 2020 (95%CI 0.42, 1.54), and a decline back to a non-significant change in January 2021 (B = 0.28, 95%CI -0.27, 1.38) compared with the pre-pandemic estimate.

None of the interactions were significant, suggesting that changes over time were unlikely to have substantially varied across groups.

## 4. DISCUSSION

This brief report presents new estimates of smoking trajectories over the first year of the pandemic in a large longitudinal sample of UK middle-aged adults. Results suggest that smoking has generally declined in this group during this period, with a meaningful portion of this change occurring by April 2020. This supports the idea that many of the succesful quit attempts that occurred at the start of the pandemic may have persisted. Results also suggest that cigarette consumption among smokers increased in April and September 2020, but decreased to reach back its pre-pandemic level in January 2021. The extent to which this may simply capture seasonal effects (i.e., higher consumption in summer months compared with winter months) needs to be investigated further.^7,8^ The results finally suggest that these changes did not meaningfully vary across socioeconomic groups. This supports the idea that, whereas inequalities in smoking remain large in the UK population, the negative consequences of the COVID-19 pandemic (which have also been unequally experienced) are unlikely to have increased inequalities in smoking during this period.

This study is not without limitations. Only 26% of those aged 35-64 with a full interview in 2018-19 were included in our analytic sample, meaning that our analyses are unlikely to be representative despite the use of the survey weights provided by the UKHLS team. The focus on middle-aged adults precludes generalisations to other age groups, particularly younger adults who are still in the process of initiating smoking^9,10^ and have faced the brunt of the economic consequences of the pandemic.^11^ Results from the English Smoking Toolkit Study, which recorded a potential increase in smoking prevalence among those aged 16-24 between 2019 and 2021, support the call for better evidence in young adults.^12^ The findings here may also not align with the reality of other countries in keeping with the intensity of the impact of the pandemic and the government’s response.^13^ Despite these concerns, the findings represent encouraging results. Since disasters and recessions affect individuals over a long period of time, studies will be needed to understand the long-term effects of the pandemic on smoking trajectories over the next decade.

## Data Availability

The UKHLS main and COVID-19 datasets can be accessed from the UK Data Service.

https://ukdataservice.ac.uk/

## ACKNOWLEDGEMENTS

TG is a Banting Postdoctoral Fellow funded by the Canadian Institutes of Health Research. TG thanks the University of Tübingen for hosting him during the writing of this brief report. UKHLS is an initiative funded by the Economic and Social Research Council and various Government Departments, with scientific leadership by the Institute for Social and Economic Research, University of Essex, and survey delivery by NatCen Social Research and Kantar Public. The research data are distributed by the UK Data Service.

## CONTRIBUTORSHIP STATEMENT

TG designed the study, accessed the data, ran the analyses, interpreted the results, and wrote the paper. He confirms his personal full access to all aspects of the research and writing process, and takes final responsibility for the paper.

