## Supplementary Material for "Smoking trajectories over the first year of the pandemic in UK middle-aged adults: evidence from the UKHLS COVID-19 study"

### Title

### Table of contents

1. Validity of smoking prevalence across age groups, by sex.
2. Smoking transitions, by sex.
3. Models for smoking prevalence, by sex.
4. Models for cigarettes smoked per day among smokers, by sex.
5. Summary of interactions.

**SUPPLEMENTARY TABLE 1**
**Prevalence of smoking across time points, 2018-21. UKHLS COVID CW7 sample.**

| | | 2018-19 | 2018-19 | Apr 2020 | Sep 2020 | Jan 2021 |
|---|---|---|---|---|---|---|
| **Age** | **Sex** | **%** | **%** | **%** | **%** | **%** |
| | | **HSE** | **Cross-sectional CW7 weight (n = 9,642)** | | | |
| **16-34** | **M** | 24.7 | 10.9 | 8.2 | 9.4 | 9.5 |
| | **F** | 19.6 | 13.1 | 10.8 | 8.9 | 10.7 |
| **35-64** | **M** | 18.9 | 15.3 | 13.5 | 14.3 | 12.5 |
| | **F** | 15.2 | 14.6 | 13.0 | 12.0 | 12.5 |
| **65+** | **M** | 7.6 | 4.6 | 4.5 | 4.0 | 4.1 |
| | **F** | 8.2 | 4.3 | 4.3 | 4.1 | 3.8 |
| | | **HSE** | **Longitudinal CW7 weight (n = 7,449)** | | | |
| **16-34** | **M** | 24.7 | 7.9 | 5.7 | 6.5 | 6.2 |
| | **F** | 19.6 | 9.9 | 9.0 | 7.8 | 8.8 |
| **35-64** | **M** | 18.9 | 15.5 | 15.1 | 15.6 | 14.2 |
| | **F** | 15.2 | 14.1 | 12.7 | 12.5 | 12.0 |
| **65+** | **M** | 7.6 | 4.4 | 4.3 | 4.0 | 4.0 |
| | **F** | 8.2 | 3.1 | 3.1 | 2.9 | 2.8 |

Sample is everyone who had a full interview in 2018-19 and had either a cross-sectional or longitudinal weight in January 2021. Estimates are weighted using the longitudinal C7 weight.

The Health Survey for England (HSE) estimate is for the English population based on the pooled samples for 2018 and 2019. England accounts for roughly 84% of the UK population.

**SUPPLEMENTARY TABLE 2**

**Smoking transitions between waves, UK adults ages 35-64. UKHLS COVID CW7 longitudinal sample (2018-21).**

| From Wave 10 to Wave C1 | | From Wave 10 to Wave C1 | | From Wave 10 to Wave C1 | |
|---|---|---|---|---|---|
| 2018-19 to April 2020 | | 2018-19 to April 2020 | | 2018-19 to April 2020 | |
| **EVERYONE** | | | | | |
| **Smokers** | | **Smokers** | | **Smokers** | |
| Stayed smoker | 83.9 | Stayed smoker | 92.6 | Stayed smoker | 88.9 |
| Became non-smoker | 16.1 | Became non-smoker | 7.4 | Became non-smoker | 11.1 |
| **Non-smokers** | | **Non-smokers** | | **Non-smokers** | |
| Became smoker | 1.7 | Became smoker | 1.3 | Became smoker | 0.8 |
| Stayed non-smoker | 98.3 | Stayed non-smoker | 98.7 | Stayed non-smoker | 99.2 |
| **MALES** | | | | | |
| **Smokers** | | **Smokers** | | **Smokers** | |
| Stayed smoker | 84.0 | Stayed smoker | 94.3 | Stayed smoker | 87.4 |
| Became non-smoker | 16.0 | Became non-smoker | 5.7 | Became non-smoker | 12.3 |
| **Non-smokers** | | **Non-smokers** | | **Non-smokers** | |
| Became smoker | 2.3 | Became smoker | 1.7 | Became smoker | 0.5 |
| Stayed non-smoker | 97.7 | Stayed non-smoker | 98.3 | Stayed non-smoker | 99.5 |
| **FEMALES** | | | | | |
| **Smokers** | | **Smokers** | | **Smokers** | |
| Stayed smoker | 83.8 | Stayed smoker | 90.9 | Stayed smoker | 90.6 |
| Became non-smoker | 16.2 | Became non-smoker | 9.1 | Became non-smoker | 9.4 |
| **Non-smokers** | | **Non-smokers** | | **Non-smokers** | |
| Became smoker | 1.1 | Became smoker | 0.9 | Became smoker | 1.1 |
| Stayed non-smoker | 98.9 | Stayed non-smoker | 99.1 | Stayed non-smoker | 98.9 |

N participants = 4,310. Estimates are weighted using the longitudinal C7 weight.

**SUPPLEMENTARY TABLE 3**

**Testing changes in <u>smoking prevalence</u> over time, UK adults ages 35-64. UKHLS COVID CW7 longitudinal sample (2018-21).**

| | **Overall** *N obs* = 17,120 | | | **Males** *N* obs = 6,814 | | | **Females** *N* obs = 10,306 | | |
|---|---|---|---|---|---|---|---|---|---|
| | **PR** | **95%CI** | ***p*** | **PR** | **95%CI** | ***p*** | **PR** | **95%CI** | ***p*** |
| | **Population-average (Pooled with clustered SEs)** | | | | | | | | |
| **Time** | | | | | | | | | |
| (ref. 2018-19) | --- | --- | --- | --- | --- | --- | --- | --- | --- |
| CW1 – April 2020 | 0.94 | 0.87-1.01 | .081 | 0.97 | 0.87-1.09 | .600 | **0.90** | **0.82-0.99** | **.037** |
| CW5 – Sept 2020 | 0.95 | 0.88-1.02 | .158 | 1.01 | 0.91-1.11 | .892 | **0.89** | **0.79-0.99** | **.045** |
| CW7 – Jan 2021 | **0.89** | **0.80-0.99** | **.027** | 0.91 | 0.77-1.08 | .287 | **0.87** | **0.77-0.97** | **.016** |
| | **Subject-specific (Random-intercept modelling)** | | | | | | | | |
| **Time** | | | | | | | | | |
| (ref. 2018-19) | --- | --- | --- | --- | --- | --- | --- | --- | --- |
| CW1 – April 2020 | **0.93** | **0.87-0.99** | **.026** | 0.96 | 0.88-1.05 | .349 | **0.91** | **0.84-0.99** | **.021** |
| CW5 – Sept 2020 | **0.92** | **0.86-0.98** | **.017** | 0.96 | 0.88-1.06 | .448 | **0.88** | **0.80-0.97** | **.007** |
| CW7 – Jan 2021 | **0.89** | **0.83-0.95** | **.001** | 0.92 | 0.84-1.02 | .107 | **0.86** | **0.79-0.94** | **.001** |

Sample is those aged 35-64 with a full interview in the UKHLS main wave 10 (2018-19) and with a valid longintudinal weight value in the COVID-19 survey wave 7 (January 2021). Estimates are weighted.

**SUPPLEMENTARY TABLE 4**
**Testing changes in cigarettes per day among smokers over time, UK adults ages 35-64. UKHLS COVID CW7 longitudinal sample (2018-21).**

| | **Overall** *N obs* **= 1,352** | | | **Males** *N* **obs = 616** | | | **Females** *N* **obs = 736** | | |
|---|---|---|---|---|---|---|---|---|---|
| | **B** | **95%CI** | ***p*** | **B** | **95%CI** | ***p*** | **B** | **95%CI** | ***p*** |
| | **Population-average (Pooled with clustered SEs)** | | | | | | | | |
| **Time** | | | | | | | | | |
| (ref. 2018-19) | --- | --- | --- | --- | --- | --- | --- | --- | --- |
| CW1 – April 2020 | 1.16 | 0.04, 2.27 | .043 | 0.91 | -0.58, 2.40 | .230 | 1.37 | -0.27, 3.00 | .102 |
| CW5 – Sept 2020 | 0.74 | -0.46, 1.94 | .225 | 1.05 | -0.27, 2.37 | .119 | 0.27 | -1.69, 2.23 | .786 |
| CW7 – Jan 2021 | 0.50 | -0.52, 1.51 | .338 | 0.88 | -0.52, 2.28 | .218 | 0.06 | -1.40, 1.52 | .938 |
| | **Subject-specific (Random-intercept modelling)** | | | | | | | | |
| **Time** | | | | | | | | | |
| (ref. 2018-19) | --- | --- | --- | --- | --- | --- | --- | --- | --- |
| CW1 – April 2020 | **0.51** | **0.03, 1.00** | **.038** | 0.14 | -0.71, 0.99 | .748 | **0.82** | **0.15, 1.49** | **.017** |
| CW5 – Sept 2020 | **0.98** | **0.42, 1.54** | **.001** | **1.03** | **0.14, 1.92** | .023 | **0.92** | **0.27, 1.57** | **.005** |
| CW7 – Jan 2021 | 0.28 | -0.27, 0.83 | .316 | 0.08 | -0.75, 0.91 | .847 | 0.43 | -0.24, 1.10 | .211 |

Sample is those aged 35-64 with a full interview in the UKHLS main wave 10 (2018-19) and with a valid longintudinal weight value in the COVID-19 survey wave 7 (January 2021). Estimates are weighted.

**SUPPLEMENTARY TABLE 5**
**Differences for changes over time. UKHLS COVID CW7 sample, 2018-21.**

| | **Smoking prevalence** | | | **Cigarettes per day** |
|---|---|---|---|---|
| | **Everyone** | **Males** | **Females** | **Smokers** |
| **Effect modifiers in 2018-19** | ***p*** | ***P*** | ***p*** | ***p*** |
| Sex | .600 | --- | --- | .401 |
| Having a “White UK” ethnicity (yes, no) | .558 | .748 | .730 | .423 |
| Living with a partner (yes, no) | .995 | .917 | .761 | .524 |
| Having a degree (yes, no) | .279 | .847 | .243 | .707 |
| NS-SEC of current job (four categories) | .398 | .576 | .537 | .579 |
| Household income (five quintiles) | .212 | .178 | .502 | .246 |

*P-values* represent joint tests of significance for differences across any of the four time points, run after random-intercept models with a set of interaction dummy terms for time.
